# Benefits of sphingosine-1-phosphate receptor modulators in relapsing MS estimated with a treatment sequence model

**DOI:** 10.1101/2022.12.23.22283885

**Authors:** Cato E.A. Corsten, Simone A. Huygens, Matthijs M. Versteegh, Beatrijs H.A. Wokke, Ide Smets, Joost Smolders

## Abstract

**Background:** Three sphingosine-1-phosphate receptor (S1PR) modulators are currently available as disease-modifying therapies (DMTs) for relapsing MS in the Netherlands (i.e. fingolimod, ozanimod and ponesimod). We aimed to identify which S1PR modulator yields the highest benefit from a health-economic and societal perspective during a patient’s lifespan.

**Methods:** Incorporating Dutch DMT list prices, we used the ErasmusMC/iMTA MS model to compare DMT sequences, including S1PR modulators and eight other DMT classes, for treatment-naive patients with relapsing MS in terms of health outcomes (number of lifetime relapses, time to Expanded Disability Status Scale (EDSS) 6, lifetime quality-adjusted life years (QALYs)) and cost-effectiveness (net health benefit (NHB)). We estimated the influence of list price and EDSS progression on cost-effectiveness outcomes.

**Results:** In deterministic and probabilistic analysis, DMT sequences with ponesimod have lower lifetime costs and higher QALYs resulting in a higher average NHB compared to sequences with other S1PR modulators. Ponesimod remains the most cost-effective S1PR modulator when EDSS progression is class-averaged. Given the variable effects on disability progression, list price reductions could make fingolimod but not ozanimod more cost-effective than ponesimod.

**Conclusion:** Our model favours ponesimod among the S1PR modulators for the treatment of relapsing MS. This implies that prioritizing ponesimod over other S1PR modulators translates into a more efficacious spending of national healthcare budget without reducing benefit for people with MS. Prioritizing cost-effective choices when counselling patients contributes to affordable and accessible MS care.

**Highlights:**

- Disease-modifying therapies (DMTs) are the largest cost driver in MS
- Three sphingosine-1-phosphate receptor (S1PR) modulators are used for relapsing MS
- Our model compares sequences of up to 5 DMTs in terms of health (relapses, progression) and costs
- Comparing sequences including S1PR modulators, ponesimod is the most cost-effective drug
- Cost-effectiveness research helps to prioritize when prescribing drugs of a similar class

## Introduction

Over the past decade, the MS treatment landscape has burgeoned and we now have disease-modifying therapies (DMTs) available from nine different drug classes.^1^ Unavoidably, the wide range of DMTs renders treatment strategies more complex and neurologists need to choose between escalation versus induction therapy, between different drug classes and between several similar DMTs per drug class. Moreover, there is a societal need to make evidence-based DMT choices given the variability in list price (i.e. publicly available DMT price as set by the manufacturer), DMTs being the largest cost driver in MS and considerable economic burden of chronic diseases such as MS.^2, 3^ Since the registration of fingolimod as an MS treatment in 2010, two other sphingosine-1-phosphate receptor (S1PR) modulators (ozanimod, ponesimod) have been approved for treatment of relapsing MS and one (siponimod) for secondary progressive MS. Although fingolimod was initially positioned as a second-line therapy in several countries, the newer compounds shifted the use of S1PR modulators towards treatment-naïve people with MS (pwMS).^1^ The S1PR modulators have a unique mechanism of action by sequestration of lymphocytes in lymph nodes. All compounds act on the S1PR1-subtype with fingolimod, ozanimod and siponimod also targeting S1PR5 and fingolimod S1PR3 and S1PR4.^4, 5^ The different S1PR binding profiles explain cardiologic side effects (S1PR1, S1PR3)^4^ and humoral response to vaccines (S1PR4)^6^ whereas animal studies and *ex vivo* data suggest a role in astrogliosis (S1PR1, S1PR3)^7^ and oligodendrocyte maturation (S1PR5).^8^ As no head-to-head trials of the S1PR modulators are available and different active comparators were used in the phase-3 trials, it is difficult to compare effectiveness for treatment of relapsing MS.^9-13^ These type of studies are also unlikely to follow due to the high costs involved and conflicting interests of pharmaceutical companies. In clinical practice, neurologists thus perceive three pathophysiological comparable S1PR modulators which mainly differ in terms of side effect profile. The choice between S1PR modulators is therefore primarily guided by practical considerations and personal preferences rather than data-driven insights. Given the considerable overlap in mechanism of action and differences in list price, we used a health-economic approach to generate new insights regarding the granularity between S1PR modulators. We aim to identify which one provides the highest benefit in terms of health (e.g. relapse rate reduction, disability prevention) and cost-effectiveness for the treatment of relapsing MS.

## Methods

### Erasmus MC/iMTA MS model

The ErasmusMC/iMTA MS model^14, 15^ was used to compare the benefits of DMT sequences including S1PR modulators for treatment-naïve relapsing pwMS in terms of health outcomes (i.e. number of lifetime relapses, time to Expanded Disability Status Scale (EDSS) 6, lifetime quality-adjusted life years (QALYs)) and cost-effectiveness (i.e. net health benefit (NHB)). The model evaluates the costs and benefits of DMT sequences rather than the lifetime use of single DMT. In the model, pwMS switch to another DMT when they experience unacceptable side effects or inflammatory disease activity and can receive up to five DMTs during their lifetime corresponding to line 1a, 1b, 2, 3 or 4 (Figure S1.1). A small proportion of pwMS in line 1a who experience intolerable side effects will switch to another first-line drug corresponding to line 1b. The model counts the costs and QALYs of pwMS during their disease course. The costs cover the DMTs, other healthcare use (i.e. DMT administration, monitoring, inpatient care, day admissions, tests, other medication, relapses, and DMT switches), productivity loss and informal care of pwMS. Since the initial publication of the ErasmusMC/iMTA MS model^14^, it has been updated to reflect the evolving DMT landscape (Supplemental Material 1). Most relevant is the use of S1PR modulators as first- and second-line treatment.

### Efficacy and costs of S1PR modulators

In the model, the efficacy (in terms of annualized relapse rates and 24-week confirmed disability progression (CDP)) of S1PR modulators (and other DMTs) was based on network meta-analyses (NMA, Supplemental Material 2) and was assumed to be constant irrespective of their position in the DMT sequence (i.e. first-versus second-line). The drug acquisition costs of DMTs were based on the most recent (2022) publicly available list prices in the Netherlands (Table 1).^16^

**Table 1.** Efficacy as estimated in the network meta-analysis and Dutch costs of S1PR modulators.

| DMT | Incidence rate ratio of ARR vs. placebo [95% CI] | Relative risk of 24-week confirmed disease progression vs. placebo [95% CI] | Annual drug acquisition costs in the Netherlands (in Euros) |  |
| --- | --- | --- | --- | --- |
|  |  |  | First year | Subsequent years |
| FIN | 0.45 [0.36-0.54] | 0.71 [0.56-0.90] | 20,591 | 20,591 |
| OZA | 0.53 [0.41-0.69] | 0.85 [0.53-1.36] | 14,936 | 15,144 |
| PON | 0.48 [0.33-0.69] | 0.64 [0.38-1.08] | 12,050 | 12,384 |
| S1PR clustered | NA | 0.71 [0.61-0.90] | NA | NA |

Abbreviations: ARR: annualized relapse rate, CI: confidence interval, DMT: disease modifying therapy, FIN: fingolimod, OZA: ozanimod, PON: ponesimod.
Details on the network meta-analysis can be found in the Supplemental Material 1 and 2. For amounts in Euro integers were used.

### DMT sequences

The following DMTs are available in the Netherlands and included in the model: interferon bèta (IFNB), dimethyl fumarate 240 mg (DMF), teriflunomide 14 mg (TER), glatiramer acetate 20 mg (GLA), ponesimod 20 mg (PON), ozanimod 1 mg (OZA), fingolimod 0.5 mg (FIN), natalizumab 300 mg (NAT), cladribine 3.5 mg/kg (CLA), ocrelizumab 600 mg (OCR), ofatumumab 20 mg (OFA), and alemtuzumab 12 mg (ALE). Glatiramer acetate 40 mg was not included because the trial only reported 12-week CDP.^17^ Further, we did not include siponimod as it is not registered for relapsing MS. In addition, PON, OZA, FIN, IFNB, TER, GLA and DMF were included in the DMT sequences as first-line treatments (line 1a and 1b), PON, OZA, FIN, NAT, CLA, OCR and OFA as second-line treatments (line 2), and NAT, CLA, OCR, OFA as third-line treatments (line 3). ALE was restricted to a last line of treatment (line 4). We only included clinically plausible DMT sequences (i.e. excluding switches within a drug class, or switching to an S1PR modulator after CLA, OCR, OFA or NAT) resulting in 504 possible DMT sequences of which 384 include an S1PR modulator (128 sequences per S1PR modulator of which 40 in line 1a and 1b, and 48 in line 2).

### Health-economic analyses

The model simulated 10,000 pwMS per DMT sequence and the outcomes represent the average of this virtual population. Based on this principle, we conducted three main analyses. First, we identified the best performing S1PR modulator in terms of cost-effectiveness (i.e. NHB), when positioned as first- or second-line in a DMT sequence. We calculated the benefits of DMT sequences with a specific DMT by averaging the outcomes of all DMT sequences with that specific DMT. Second, we performed a sensitivity analysis where we clustered the S1PR modulators in the NMA regarding their effect on 24-week CDP and applied the average in the model. This analysis was done to address uncertainty in estimation of EDSS progression as RCTs were only powered to estimate annualised relapse rate. Third, we estimated the required price reduction for the two other S1PR modulators to replace the S1PR modulator of the most cost-effective DMT sequence in first- or second-line. In accordance with the Dutch guideline for conducting economic evaluations in healthcare^18^, the analyses were performed from a societal perspective (i.e. including healthcare costs and societal costs). Costs and QALYs were discounted with 4% and 1.5%, respectively, to account for the diminishing value of future costs and benefits. ^18^ All costs in this paper represent lifetime net present costs.

To assess the uncertainty of our outcomes, all model parameters (such as efficacy and cost estimates of DMTs) were varied simultaneously by sampling from their distributions in a probabilistic sensitivity analysis (PSA). The PSA was performed for three DMT sequences starting with fingolimod, ozanimod or ponesimod and was conducted with 500 sampling iterations, while sampling 1,000 pwMS per DMT sequence.

### Definitions

*Utility* reflects how desirable or undesirable it is to live in a certain state of health. Utility as used in this model is influenced by the EDSS score^19^, relapses^20^, and adverse events (e.g. progressive multifocal leukoencephalopathy^21^, auto-immune thyroid events^22^). Utility ranges between 1 (best health) and some negative value (worst health, that can be worse than death) at the lower end with 0 representing death.

*QALY* is single metric expressing both length and quality of life (i.e., utility). It is obtained by multiplying the time spent in some state of health (such as the period spent in a certain EDSS class) with its utility value. For example, living for two years with EDSS score 2 without a relapse would yield 0.782*2 = 1.564 QALY, while the same period of two years in perfect health would yield 2 QALY. In this paper, lifetime QALYs reflect ‘clinical effectiveness’.

*Net health benefit (NHB)* reflects the cost-effectiveness of a treatment and expresses the net benefit of a treatment in terms of QALYs, adjusted for the cost given the value of a QALY. To calculate the NHB, the total lifetime costs are divided by the value of a QALY (the threshold value), hence expressing costs in terms of QALYs. These are then subtracted from total lifetime QALYs: NHB = QALYs – (costs/cost-per-QALY threshold). The appropriate cost-per-QALY threshold for pwMS in the Netherlands is € 50,000/QALY.^14, 23^

## Results

### 1. Cost-effectiveness of S1PR modulators as first- and second-line treatment

In our model, both in line 1a and line 2, ponesimod has lower lifetime costs and higher QALYs resulting in a higher average NHB of DMT sequences with ponesimod compared to sequences with ozanimod or fingolimod (Table 2). In first-line, ponesimod on average saves €62,354 compared to fingolimod and €43,850 compared to ozanimod per patient over an individual’s lifetime. In second-line, ponesimod saves €55,798 versus fingolimod and €17,472 versus ozanimod. The higher number of QALYs is explained by the longer time to EDSS 6 of ponesimod compared to the other S1PR modulators. The number of lifetime relapses is marginally higher with ponesimod than with the other S1PR modulators (Table 2). Although first-line ponesimod is associated with higher costs than second-line ponesimod, the number of QALYs is also higher resulting in a higher NHB. This implies that use of ponesimod as a first-line treatment is more cost-effective than as a second-line treatment (Table 2). Figure 1 shows that despite considerable uncertainty ponesimod is most likely to be more cost-effective than the other S1PR modulators in line 1. This translates into a probability of ponesimod being cost-effective compared to fingolimod or ozanimod at the current list prices and at a willingness-to-pay of €50,000/QALY of 90.2% and 95.4%, respectively. This means that 90.2% or 95.4% of the iterations fall above the reference line of the willingness-to-pay threshold.

**Table 2.**
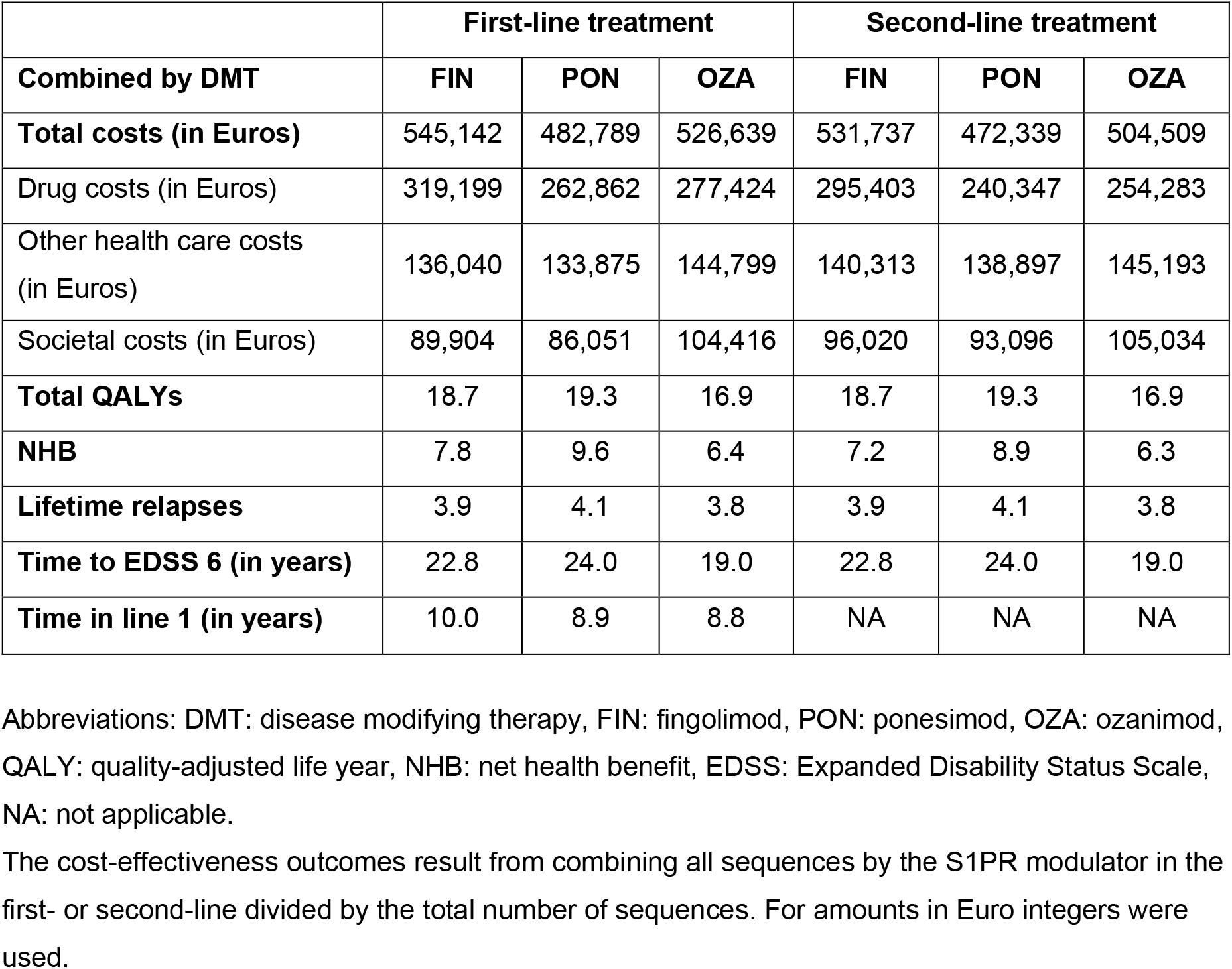
Mean cost-effectiveness outcomes of S1PR modulators.

**Figure 1.**
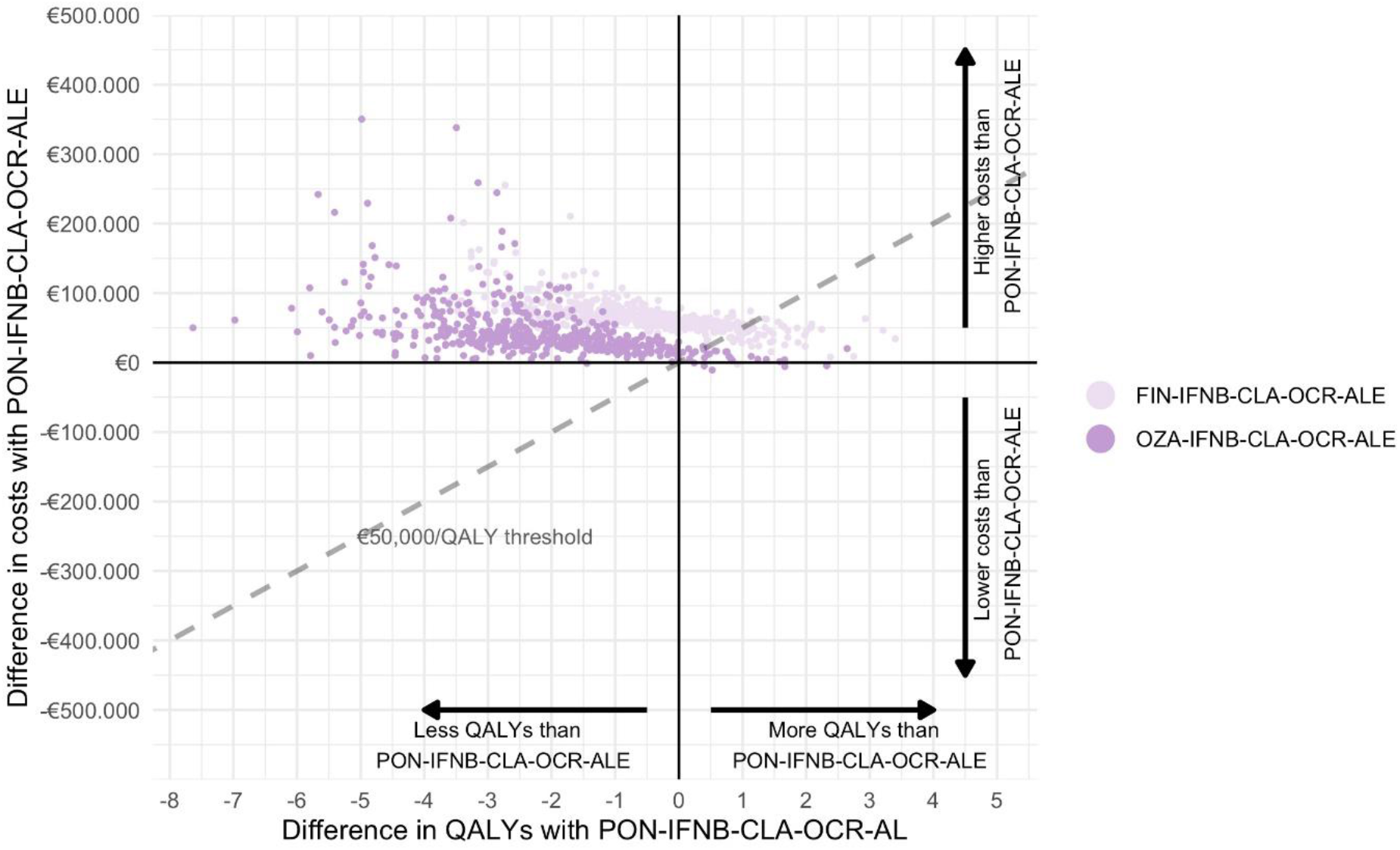
Probabilistic sensitivity analysis of the most cost-effective S1PR modulator sequences. Incremental cost-effectiveness plane of DMT sequences starting with fingolimod (FIN-IFNB-CLA-OCR-ALE, light purple) or ozanimod (OZA-IFNB-CLA-OCR-ALE, dark purple) compared to ponesimod (PON-IFNB-CLA-OCR-ALE) illustrating the uncertainty of the results. The dashed line represents the willingness-to-pay threshold of €50,000/QALY. Abbreviations: PON: ponesimod, FIN: fingolimod, OZA: ozanimod, IFNB: interferon beta, CLA: cladribine, OCR: ocrelizumab, ALE: alemtuzumab

### 2. Cost-effectiveness of S1PR modulators with class-averaged EDSS progression

If we assume that there is no difference in the efficacy of S1PR modulators to slow down EDSS progression, ponesimod remains the most cost-effective first- or second-line treatment in our model (Table 3). In this analysis, ozanimod has a higher NHB than fingolimod and thus becomes more cost-effective.

**Table 3.** Mean cost-effectiveness outcomes of S1PR modulators using a class-averaged EDSS progression.

|  | **First-line treatment** |  |  | **Second-line treatment** |  |  |
| --- | --- | --- | --- | --- | --- | --- |
| **Combined by DMT** | **FIN** | **PON** | **OZA** | **FIN** | **PON** | **OZA** |
| **Total costs (in Euros)** | 547,750 | 491,096 | 507,268 | 532,687 | 476,890 | 494,362 |
| Drug costs (in Euros) | 319,094 | 263,165 | 278,083 | 294,683 | 239,677 | 256,796 |
| Other health care costs (in Euros) | 137,084 | 136,959 | 137,211 | 140,900 | 140,727 | 140,709 |
| Societal costs (in Euros) | 91,572 | 90,972 | 91,974 | 97,103 | 96,486 | 96,857 |
| **Total QALYs** | 18.5 | 18.7 | 18.5 | 17.7 | 17.8 | 17.7 |
| **NHB** | 7.6 | 8.8 | 8.3 | 7.0 | 8.3 | 7.9 |
| **Lifetime relapses** | 3.9 | 4.0 | 3.8 | 4.5 | 4.6 | 4.4 |
| **Time to EDSS 6 (in years)** | 22.3 | 22.6 | 22.2 | 20.5 | 20.8 | 20.6 |
| **Time in line 1 (in years)** | 9.9 | 8.6 | 9.6 | NA | NA | NA |

Abbreviations: DMT: disease modifying therapy. FIN: fingolimod, PON: ponesimod, OZA: ozanimod, QALY: quality-adjusted life year, NHB: net health benefit, EDSS: Expanded Disability Status Scale, NA: not applicable
The cost-effectiveness outcomes result from combining all sequences by the S1PR modulator in the first- or second-line divided by the total number of sequences. For amounts in Euro integers were used.

### 3. Cost-effectiveness of S1PR modulators at a reduced generic price

In our model, the most cost-effective sequences with ponesimod in the first or second line are PON-IFNB-CLA-OCR-ALE and IFNB-GLA-PON-CLA-ALE. Fingolimod’s price should be 63% lower than its 2022 list price (i.e. a maximum of €20.87 daily) to replace ponesimod in these sequences. At that price, prescribing fingolimod instead of ponesimod in line 1 or 2 would on average save €27,171 or €23,058 per patient per lifetime, respectively. These price reductions are, however, contingent on the price of ponesimod. For each 10% price decline of ponesimod (in either first- or second-line), an additional price decline of fingolimod of about 5% is necessary. As even a zero price for ozanimod could not offset the costs associated with the lower amount of QALYs with ozanimod compared to ponesimod, there is no price reduction that would render ozanimod more cost-effective than ponesimod.

## Discussion

Using the Erasmus MC/iMTA MS model, ponesimod is modelled to be the most cost-effective S1PR modulator as a first- or second-line treatment of relapsing MS at current list prices. This implies that prioritizing ponesimod over other S1PR modulators can result in a more efficacious spending of national healthcare budget. Treatment sequences with ponesimod namely surpass sequences with ozanimod and fingolimod in terms of QALYs and NHB, explained by longer time to EDSS 6 in ponesimod. Although a slightly higher number of relapses is seen in ponesimod compared to the other S1PR modulators, this observation does not offset the overall benefits from a societal-economic perspective due to the limited impact on quality of life and relapse-associated healthcare costs versus disability accrual. Although our model builds on certain assumptions about the comparability of S1PR modulators, the probabilistic analysis shows that our main conclusions hold when this uncertainty is modelled.

The cost-effectiveness of ponesimod among the S1PR modulators in our model is highly driven by its superior effect on EDSS progression in the NMA compared to fingolimod and especially ozanimod. As no head-to-head comparisons exist and as there is no biological underpinning of this observation^4^, it is impossible to exclude whether this discrepancy reflects differences in patient characteristics between phase-3 trials rather than true superiority of ponesimod when it comes to disability prevention. Furthermore, the NMA shows the difference to be uncertain. Differences in patient demographics have namely been known to influence efficacy of fingolimod on disability progression, with negative results in a phase-3 trial in primary progressive MS.^24^ At first sight, the patient characteristics between the ozanimod (RADIANCE, SUNBEAM) and ponesimod (OPTIMUM) phase-3 trial look similar in terms of baseline EDSS and number of relapses previous to inclusion. However, much less details are provided on the pwMS included in the ozanimod trials (e.g. number of pwMS fulfilling criteria for highly active disease) making it impossible to assess whether they represented a more benign population.^11-13^ In addition, people from Eastern Europe were slightly overrepresented in RADIANCE and SUNBEAM compared to OPTIMUM and especially the pivotal fingolimod trials (FREEDOMS I&II).^25^ This might be relevant as lower rates of EDSS progression have been reported in placebo data from randomized trials in Eastern European countries (10.8%) vs. Western Europe (13.1%) and the USA/Canada (23.1%) which could not be explained by differences of baseline characteristics.^26^ Another consideration is the shorter follow-up duration of SUNBEAM (i.e. 12 months) which potentially reduced statistical power to detect meaningful differences in disability progression.^12^ All of the above are however of unknown significance, and cannot obliterate the fact that the results on 24-week CDP of ozanimod were confirmed by two independent phase-3 trials including over 800 pwMS in the treatment arm.^11, 12^ Moreover, our data shows that even when pooling the effect on 24-week CDP, ponesimod remains the most cost-effective choice when treating pwMS with an S1PR modulator. In the model, this is now mainly driven by the favourable list price of ponesimod compared to ozanimod and fingolimod and thus prone to change. However, in clinical practice, where health economic evidence to date sparsely drives decisions, ponesimod’s pharmacodynamics and side effect profile can add to economic considerations.

Cost-effectiveness research is a key instrument to keep our healthcare affordable, especially in the context of chronic diseases such as MS requiring long-term treatment.^27^ The economic burden of MS is mainly driven by its direct medical costs ^28^, with DMTs covering more than half of the total medical costs per patient.^2^ In our model, the most recent DMT acquisition costs were used as they are publicly available in the Netherlands, which is essential to conduct this type of research.^16^ Although parallel trade of DMTs and price negotiations between private pharmacies and pharmacological companies are common practice in the Netherlands, these practices will not reduce applicability of the model when these rebates are local savings that do not impact on the amount reimbursed by the Dutch health insurance. The societal expenses for these DMTs remain the same regardless of whether their profitability is high for the manufacturer or for the hospital in which it is prescribed. As significant differences in pricing have been documented among the EU member states with eleven-fold differences in relation to interferon-beta between Germany and Croatia^29^, our findings cannot be automatically generalised to other countries. However, this would only apply to differences between cost-effectiveness of ponesimod versus fingolimod as the lower quality of life generated by ozanimod could never result in a cost-effective DMT choice. Due to the loss of exclusivity of fingolimod, the estimation of the required reduction for (generic) fingolimod to be the most cost-effective first- or second-line S1PR modulator, would need to be at least 63%. Median price declines for generics have been shown to be 41% four years after introduction in The Netherlands.^30^ Variability in these reductions are, however, large and hence it is not possible to state if these price reductions are achieved for fingolimod and, more importantly, if such reductions translate into societal savings rather than local savings.

An important limitation of our study is that the model only considers prescriber preferences and does not take into account patient-specific factors such as preferences regarding mode of administration, likelihood of a humoral response to vaccines and family planning. Undoubtedly, these factors play an important role in the decision-making process. Especially the fact that all S1PR modulators are contra-indicated during conception, pregnancy and lactation^31^ could hamper prioritizing cost-effective choices in daily practice. Second, the impact of rebound disease activity after S1PR cessation, particularly of fingolimod, on disability accrual and healthcare expenses, was not taken into account.^32, 33^ Along this line, we included the treatment initiation costs of fingolimod to mitigate its cardiac side effects but no other healthcare and S1PR modulator-specific monitoring costs. Consequences of side effects were restricted to switching treatment and subsequent healthcare costs, as explained in our previous work.^14^ Importantly, although broader treatment sequences were previously analysed^15^, we did not assess the cost-effectiveness of sequences with S1PR modulators in first- or second-line relative to sequences with other DMTs in this paper. Furthermore, although list prices are publicly available, there is a widespread lack of transparency regarding true DMT costs. Price reductions following negotiations between pharmacies, hospitals and insurance companies remain typically undisclosed. Our insights regarding the cost-effectiveness of S1PR modulators should incentivize all involved parties to transfer any local savings into societal savings. On a wider note, cost-effectiveness of escalation strategies with S1PR modulators or other DMTs should be weighed against induction strategies with or without immune reconstitution, which is a topic of further research. Finally, the results of this study for the Dutch setting may not be transferable to other countries due to country-specific list prices of DMTs and switching practices. Although the health economic model settings might be specific to The Netherlands, the principles of weighing benefits of treatments against their costs are not.

In summary, cost-effectiveness research is a useful tool to prioritize choices when DMTs of the same drug class are available. In our model based on the Dutch context, ponesimod is the most cost-effective choice among the S1PR modulators allowing to maximize patient and financial benefits. Given these results, clinicians have a responsibility in prioritizing cost-effective choices when counselling pwMS. Combining a health-economic rationale with the principles of early treatment in MS will reduce disability accumulation while keeping MS care affordable and improve accessibility to innovative, unexplored drug classes.

## Data Availability

All data produced in the present study are available upon reasonable request to the authors

## Abbreviations

ALE: alemtuzumab
CDP: confirmed disability progression
CLA: cladribine
DMF: dimethyl fumarate
DMT: disease-modifying therapy
EDSS: Expanded Disability Status Scale
FIN: fingolimod
GLA: glatiramer acetate
IFNB: interferon bèta
IRT: immune reconstitution therapy
NAT: natalizumab
NEDA: no evidence of disease activity
NHB: net health benefit
NMA: network meta-analyses
OCR: ocrelizumab
OFA: ofatumumab
OZA: ozanimod
PON: ponesimod
PSA: probabilistic sensitivity analysis
pwMS: people with multiple sclerosis
QALY: lifetime quality-adjusted life years
S1PR: sphingosine-1-phosphate receptor
TER: teriflunomide

## Acknowledgements

None

## Funding acknowledgements

C.E.A.C. has received funding from the Dutch National MS Foundation.

S.A.H. has received funding from Merck for MS-related research.

M.M.V. has received funding from Merck for MS-related research.

