## Supplemental Material 1 for "Benefits of sphingosine-1-phosphate receptor modulators in relapsing MS estimated with a treatment sequence model"

**Supplemental material 1 - Update of ErasmusMC/iMTA MS treatment sequence model**

Since the development of the model as described in Huygens & Versteegh (2021) [1], several parts of the model have been updated to reflect the evolving disease modifying therapy (DMT) landscape.

Included DMTs

In the updated model, we now included the DMTs that have recently become available (ofatumumab, ozanimod, ponesimod, ublituximab, and rituximab), clustered interferon treatments and included fingolimod as a first-line treatment instead of only as a second-line treatment.

DMT acquisition costs

We used the most recent (2022) DMT acquisition costs based on publicly available list prices in the Netherlands (Table S1.1).[2]

**Table S1.1** Updated costs of DMTs in the Netherlands (2022 Euros)

| DMT | **Costs year 1** In Euros | **Costs year 2+** In Euros |
|---|---|---|
| **Alemtuzumab** | 34,729 | 20,837 |
| **Cladribine** | 25,890 | 25,890 |
| **Dimethyl fumarate** | 14,026 | 14,162 |
| **Fingolimod** | 20,591 | 20,591 |
| **Glatiramer acetate** | 8,784 | 8,784 |
| **Natalizumab** | 16,446 | 16,446 |
| **Ocrelizumab** | 22,437 | 22,437 |
| **Peginterferon** | 11,715 | 11,826 |
| **Teriflunomide** | 11,435 | 11,435 |
| **Interferon-beta 30** | 8,476 | 8,728 |
| **Interferon-beta 44** | 9,887 | 10,408 |
| **Interferon-beta 250** | 7,231 | 7,413 |
| **Ofatumumab** | 23,434 | 20,086 |
| **Ozanimod** | 14,936 | 15,144 |
| **Ponesimod** | 12,050 | 12,384 |
| **Rituximab** | 3,802 | 2,535 |

DMT: disease modifying treatment.

Network meta-analysis

In the network meta-analyses (NMA) of annualized relapse rates (ARR) and disability progression we now included the DMTs that have recently become available (ofatumumab, ozanimod, ponesimod, ublituximab, and rituximab), excluded daclizumab and clustered interferon treatments. In addition, we replaced 12-week confirmed disability progression (CDP) results with the more robust 24-week CDP where possible and excluded studies that did not report 12-week CDP from the disability progression NMA. The updated NMA includes 24-week CDP data for all available DMTs, except for glatiramer acetate 40 mg. More information about the methods and results of the NMA of ARR and disability progression are available in Supplementary Material 2.

The DMT-specific risk ratios of discontinuation due to adverse events were based on the NMA of Liu et al. (2021) [3] or, in case of interferons, on the outcomes of the NMA performed by the Dutch Association for Neurology. [4] In line with a previous study, the risk ratio of discontinuation due to AEs for rituximab was assumed to be equal to the risk ratio of ocrelizumab.[3]

Age-dependency of background relapse risk

The background relapse risk was based on a meta-analysis of post-2005 RCTs (ARR = 0.44 at age 37). [1] As the annualized relapse rate is known to decrease by age [5-7], we corrected this value upward for ages under 37 and downward for ages over 37. In the updated model, we used the study of Tremlett et al. (2009) for the age-dependency of relapses (i.e. ARR decreases with 22.9% per 5 years up to age 40 years and 30.5% per 5 years age >40 years).[6]

Retreatment with alemtuzumab

Retreatment with alemtuzumab was included in case of disease activity that initiates a switch according to the model's clinical decision rules[1] in pwMS whose first administration of alemtuzumab started at least 4 years ago. [8]

Auto-immune thyroid events with alemtuzumab

The risk of auto-immune thyroid events (ATE) in pwMS on alemtuzumab and their associated healthcare costs and quality of life impact were included in the model. The probability of ATE (0.33, 95% confidence interval (CI): 0.24-0.43) was based on a systematic review and meta-analysis of ATE in pwMS on alemtuzumab. [9] The majority of ATE (63%) entailed Graves' disease and therefore ATE treatment costs were based on treatments for Graves' disease. [9] After alemtuzumab treatment, 12% of the pwMS with Graves' disease remitted spontaneous while 88% of pwMS were treated with oral antithyroid drugs of which 22% were subsequently treated with radioactive iodine (RAI) and 11% underwent a total thyroidectomy. [9] The costs of antithyroid drugs (thiamazole 30 mg and levothyroxine 150 mcg) were 68 euros per year. [2] The costs of radioiodine treatment was 1,937 euro based on the weighted average of the costs of radioiodine treatment diagnostic-related groups (DRG; 049899003 and 049899004) used in 2019 in the Netherlands, including 20% re-treatment. [9] The costs of a total thyroidectomy was 2,954 euros and was also based on the costs of a DRG (049899005).The disutility of ATE was based on Donovan et al. (2016) [10] and was 0.05 in the first year of the ATE and 0.02 in subsequent years.

### Supplemental figures

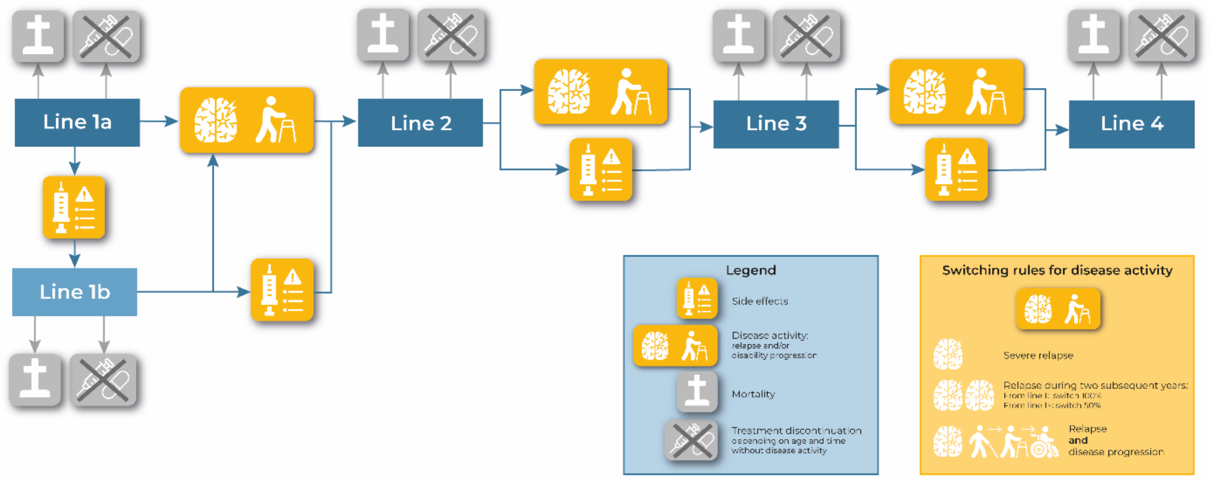


**Figure S1.1** Clinical decision rules in the ErasmusMC/iMTA model
