## Supplemental Material 2 for "Benefits of sphingosine-1-phosphate receptor modulators in relapsing MS estimated with a treatment sequence model"

**Supplemental material 2 – Methods and results of network meta-analyses**

### Introduction

Novel disease modifying therapies (DMTs) for MS have generally been tested in randomized controlled trials (RCTs) against placebo and hence head-to-head efficacy studies are not available. Therefore, we performed a network meta-analysis enabling head-to-head comparisons of all DMTs.

### Methods

Input data

The included RCTs and extracted data were derived from a recent systematic review of the US Institute for Clinical and Economic Review (US-ICER)[1,2] supplemented with an RCT of cladribine versus placebo (Table S2.1 and S2.2)[3]. We refer to chapter 4 of the US-ICER report for the search strategy, study selection, data extraction, for the quality assessment and an assessment of level of certainty in evidence.[2]

**Update 2022:** In 2022, we updated the NMAs to include the DMTs that have recently become available (i.e. ofatumumab, ozanimod, ponesimod, ublituximab, and rituximab)[4–10]. We also clustered the interferon treatments and therefore we excluded studies that only compared interferons compared to each other or placebo.[11–15] In addition, we performed an additional search to include 24-week confirmed disease progression (CDP) instead of 12-week CDP, in line with the guideline of the European Medicines Agency (EMA).[16] For six trials (including peg-interferon, teriflunomide, dimethyl fumarate and glatiramer, dimethyl fumarate, natalizumab and cladribine), we replaced the 12-week CDP data for 24-week CDP data (see references below Table S2.2). There were five studies that did not report 24-week CDP[17–21] and therefore these studies were excluded. The updated NMA includes 24-week CDP data for all available DMTs, except for glatiramer 40 mg. We performed a sensitivity analysis including the studies that only reported 12-week CDP, but this did not have a large impact on the results (see Table S2.4). Finally, we excluded two studies on daclizumab because this DMT was withdrawn from the market.[22,23]

Analyses

A head-to-head treatment efficacy was estimated in a frequentist random effects NMA for RCT endpoints on annualized relapse rates and disability progression as captured with the Expanded Disability Status Scale (EDSS) score using the R package ‘netmeta’ (version 1.2-0).[24]

Between study variance was assessed with the I-squared statistic and confidence intervals were computed for use in probabilistic sensitivity analyses of the cost-utility analysis. Direct and indirect evidence were compared using the back-calculation method in the ‘netsplit’ function of the ‘netmeta’ package.

**Table S2.1** Input data for annualized relapse rate NMA

| Reference | Study name | Year | Treatments | Sample size | Person years | Number of events |
|---|---|---|---|---|---|---|
| Jacobs 1996 | | 1996 | IFN B-1a 30 mcg IM | 158 | 293 | 196 |
| Jacobs 1996 | | 1996 | Placebo | 143 | 274 | 225 |
| Lublin 2013 | CombiRx | 2013 | IFN B-1a 30 mcg IM | 250 | 604,4 | 97 |
| Lublin 2013 | CombiRx | 2013 | Glatiramer 20 mg SC | 259 | 650,7 | 70 |
| Vollmer 2014 | BRAVO | 2014 | IFN B-1a 30 mcg IM | 447 | 825 | 215 |
| Vollmer 2014 | BRAVO | 2014 | Placebo | 450 | 809 | 275 |
| IFNB Multiple Sclerosis Study Group 1993 | | 1993 | IFN B-1b 250 mcg SC | 124 | 207 | 173 |
| IFNB Multiple Sclerosis Study Group 1993 | | 1993 | Placebo | 123 | 209.2 | 266 |
| Cadavid 2009 | BECOME | 2009 | IFN B-1b 250 mcg SC | 36 | 68.04 | 25 |
| Cadavid 2009 | BECOME | 2009 | Glatiramer 20 mg SC | 39 | 70.59 | 23 |
| O’Connor 2009 | BEYOND | 2009 | IFN B-1b 250 mcg SC | 897 | 2260 | 814 |
| O’Connor 2009 | BEYOND | 2009 | Glatiramer 20 mg SC | 448 | 1099.5 | 374 |
| Bornstein 1987 | | 1987 | Glatiramer 20 mg SC | 25 | 47.5 | 16 |
| Bornstein 1987 | | 1987 | Placebo | 23 | 45.1 | 62 |
| Johnson 1995 | | 1995 | Glatiramer 20 mg SC | 125 | 273 | 161 |
| Johnson 1995 | | 1995 | Placebo | 126 | 250 | 210 |
| Khan 2013 | GALA | 2013 | Glatiramer 40 mg SC | 943 | 884.4 | 293 |
| Khan 2013 | GALA | 2013 | Placebo | 461 | 442.5 | 223 |
| Mikol 2008 | REGARD | 2008 | IFN B-1a 44 mcg SC | 386 | 669.5 | 201 |
| Mikol 2008 | REGARD | 2008 | Glatiramer 20 mg SC | 378 | 669.5 | 194 |
| Calabresi 2014 | ADVANCE | 2014 | PEG B-1a 125 mcg SC | 512 | 404.3 | 103 |
| Calabresi 2014 | ADVANCE | 2014 | Placebo | 500 | 420.9 | 167 |
| Cohen 2010 | TRANSFORMS | 2010 | Fingolimod 0,5 mg PO | 429 | 424.6 | 68 |
| Cohen 2010 | TRANSFORMS | 2010 | IFN B-1a 30 mcg IM | 431 | 415.7 | 137 |
| Kappos 2010 | FREEDOMS | 2010 | Fingolimod 0,5 mg PO | 425 | 810.3 | 146 |

| Reference | Study name | Year | Treatments | Sample size | Person years | Number of events |
|---|---|---|---|---|---|---|
| Kappos 2010 | FREEDOMS | 2010 | Placebo | 418 | 766.3 | 307 |
| Calabresi 2014 | FREEDOMS II | 2014 | Fingolimod 0,5 mg PO | 358 | 623.8 | 131 |
| Calabresi 2014 | FREEDOMS II | 2014 | Placebo | 355 | 615 | 246 |
| O’Connor 2011 | TEMSO | 2011 | Teriflunomide 7 mg PO | 365 | 633.7 | 233 |
| O’Connor 2011 | TEMSO | 2011 | Teriflunomide 14 mg PO | 358 | 615 | 227 |
| O’Connor 2011 | TEMSO | 2011 | Placebo | 363 | 627.7 | 335 |
| Confavreux 2014 | TOWER | 2014 | Teriflunomide 7 mg PO | 407 | 614 | 235 |
| Confavreux 2014 | TOWER | 2014 | Teriflunomide 14 mg PO | 370 | 573.6 | 177 |
| Confavreux 2014 | TOWER | 2014 | Placebo | 388 | 608.4 | 296 |
| Vermersch 2014 | TENERE | 2014 | Teriflunomide 7 mg PO | 109 | 136.2 | 58 |
| Vermersch 2014 | TENERE | 2014 | Teriflunomide 14 mg PO | 111 | 132.2 | 35 |
| Vermersch 2014 | TENERE | 2014 | IFN B-1a 44 mcg SC | 104 | 112.1 | 25 |
| Fox 2012 | CONFIRM | 2012 | Dimethyl fumarate 240 mg PO | 359 | 552.99 | 124 |
| Fox 2012 | CONFIRM | 2012 | Glatiramer 20 mg SC | 350 | 569.62 | 163 |
| Fox 2012 | CONFIRM | 2012 | Placebo | 363 | 561.43 | 212 |
| Gold 2012 | DEFINE | 2012 | Dimethyl fumarate 240 mg PO | 410 | 628.61 | 128 |
| Gold 2012 | DEFINE | 2012 | Placebo | 408 | 612.35 | 246 |
| Polman 2006 | AFFIRM | 2006 | Natalizumab 300 mg IV | 627 | 1200 | 276 |
| Polman 2006 | AFFIRM | 2006 | Placebo | 315 | 578 | 422 |
| Coles 2008 | CAMMS223 | 2008 | Alemtuzumab 12 mg IV | 112 | 309.09 | 34 |
| Coles 2008 | CAMMS223 | 2008 | IFN B-1a 44 mcg SC | 111 | 247.22 | 89 |
| Cohen 2012 | CARE-MS I | 2012 | Alemtuzumab 12 mg IV | 376 | 661.11 | 119 |
| Cohen 2012 | CARE-MS I | 2012 | IFN B-1a 44 mcg SC | 187 | 312.82 | 122 |
| Coles 2012 | CARE-MS II | 2012 | Alemtuzumab 12 mg IV | 426 | 907.69 | 236 |
| Coles 2012 | CARE-MS II | 2012 | IFN B-1a 44 mcg SC | 202 | 386.54 | 201 |
| Hauser 2008 | HERMES | 2008 | Rituximab 1000 mg IV | 69 | 59.2 | 21 |

| Reference | Study name | Year | Treatments | Sample size | Person years | Number of events |
|---|---|---|---|---|---|---|
| Hauser 2008 | HERMES | 2008 | Placebo | 35 | 27.2 | 19 |
| Hauser 2017 | OPERA I | 2017 | Ocrelizumab 600 mg IV | 410 | 754.3 | 121 |
| Hauser 2017 | OPERA I | 2017 | IFN B-1a 44 mcg SC | 411 | 756.2 | 219 |
| Hauser 2017 | OPERA II | 2017 | Ocrelizumab 600 mg IV | 417 | 767.2 | 123 |
| Hauser 2017 | OPERA II | 2017 | IFN B-1a 44 mcg SC | 418 | 769.1 | 223 |
| Giovannoni 2010 | CLARITY | 2010 | Placebo | 437 | 806.76 | 254 |
| Giovannoni 2010 | CLARITY | 2010 | Cladibrine 3.5 mg | 433 | 799.4 | 112 |
| Giovannoni 2010 | CLARITY | 2010 | Cladibrine 5.25 mg | 456 | 841.8 | 126 |
| Kappos 2021 | OPTIMUM | 2021 | Ponesimod 20 mg | 567 | 1178 | 242 |
| Kappos 2021 | OPTIMUM | 2021 | Teriflunomide 14 mg PO | 566 | 1176 | 344 |
| Cohen 2019 | RADIANCE | 2019 | IFN B-1a 30 mcg IM | 441 | 882 | 247 |
| Cohen 2019 | RADIANCE | 2019 | Ozanimod 0.5 mg | 439 | 878 | 193 |
| Cohen 2019 | RADIANCE | 2019 | Ozanimod 1 mg | 433 | 866 | 147 |
| Comi 2019 | SUNBEAM | 2019 | IFN B-1a 30 mcg IM | 448 | 448 | 157 |
| Comi 2019 | SUNBEAM | 2019 | Ozanimod 0.5 mg | 451 | 451 | 108 |
| Comi 2019 | SUNBEAM | 2019 | Ozanimod 1 mg | 447 | 447 | 80 |
| Hauser 2020 | ASCLEPIOS I | 2020 | Ofatumumab | 454 | 818 | 90 |
| Hauser 2020 | ASCLEPIOS I | 2020 | Teriflunomide | 452 | 805 | 177 |
| Hauser 2020 | ASCLEPIOS II | 2020 | Ofatumumab | 469 | 950 | 95 |
| Hauser 2020 | ASCLEPIOS II | 2020 | Teriflunomide | 469 | 792 | 198 |
| Steinman 2022 | ULTIMATE I | 2022 | Teriflunomide 14 mg PO | 274 | 506 | 95 |
| Steinman 2022 | ULTIMATE I | 2022 | Ublituximab IV | 271 | 500 | 38 |
| Steinman 2022 | ULTIMATE II | 2022 | Teriflunomide 14 mg PO | 272 | 502 | 89 |
| Steinman 2022 | ULTIMATE II | 2022 | Ublituximab IV | 272 | 502 | 46 |
| Svenningsson 2022 | RIFUND-MS | 2022 | Rituximab 1000 mg IV | 98 | 196 | 3 |
| Svenningsson 2022 | RIFUND-MS | 2022 | Dimethyl fumarate 240 mg PO | 97 | 194 | 16 |

**Table S2.2 Input data for disability progression NMA**

*Note: when 24 week results were not available the study was excluded from the main NMA.

| Reference | Study name | Treatments | Sample size | EDSS progression 24 weeks | EDSS progression 12 weeks | EDSS progression included in NMA* |
|---|---|---|---|---|---|---|
| Jacobs 1996 | | IFN B-1a 30 mcg IM | 158 | 35 | | 35 |
| Jacobs 1996 | | Placebo | 143 | 50 | | 50 |
| Lublin 2013 | CombiRx | IFN B-1a 30 mcg IM | 241 | 52 | | 52 |
| Lublin 2013 | CombiRx | Glatiramer 20 mg SC | 246 | 61 | | 61 |
| Vollmer 2014 | BRAVO | IFN B-1a 30 mcg IM | 447 | 35 | 47 | 35 |
| Vollmer 2014 | BRAVO | Placebo | 450 | 46 | 60 | 46 |
| IFNB MS Study Group 1993 | | IFN B-1b 250 mcg SC | 122 | 43 | | 43 |
| IFNB MS Study Group 1993 | | Placebo | 122 | 56 | | 56 |
| O'Connor 2009 | BEYOND | IFN B-1b 250 mcg SC | 897 | | 188 | not included |
| O'Connor 2009 | BEYOND | Glatiramer 20 mg SC | 448 | | 90 | not included |
| Johnson 1995 | | Glatiramer 20 mg SC | 125 | | 27 | not included |
| Johnson 1995 | | Placebo | 126 | | 31 | not included |
| Khan 2013 | GALA | Glatiramer 40 mg SC | 943 | | 42 | not included |
| Khan 2013 | GALA | Placebo | 461 | | 17 | not included |
| Mikol 2008 | REGARD | IFN B-1a 44 mcg SC | 386 | 45 | | 45 |
| Mikol 2008 | REGARD | Glatiramer 20 mg SC | 378 | 33 | | 33 |
| Calabresi 2014 | ADVANCE | PEG B-1a 125 mcg SC | 512 | 20[a] | 31 | 20 |
| Calabresi 2014 | ADVANCE | Placebo | 500 | 42[a] | 50 | 42 |
| Cohen 2010 | TRANSFORMS | Fingolimod 0,5 mg PO | 429 | | 25 | not included |
| Cohen 2010 | TRANSFORMS | IFN B-1a 30 mcg IM | 431 | | 34 | not included |
| Kappos 2010 | FREEDOMS | Fingolimod 0,5 mg PO | 425 | 53 | 75 | 53 |
| Kappos 2010 | FREEDOMS | Placebo | 418 | 79 | 101 | 79 |
| Calabresi 2014 | FREEDOMS II | Fingolimod 0,5 mg PO | 358 | 49 | 91 | 49 |
| Calabresi 2014 | FREEDOMS II | Placebo | 355 | 63 | 103 | 63 |

| Reference | Study name | Treatments | Sample size | EDSS progression 24 weeks | EDSS progression 12 weeks | EDSS progression included in NMA* |
|---|---|---|---|---|---|---|
| O'Connor 2011 | TEMSO | Teriflunomide 7 mg PO | 365 | | 68 | not included |
| O'Connor 2011 | TEMSO | Teriflunomide 14 mg PO | 358 | | 62 | not included |
| O'Connor 2011 | TEMSO | Placebo | 363 | | 86 | not included |
| Confavreux 2014 | TOWER | Teriflunomide 7 mg PO | 407 | 44[b] | 68[b] | 44 |
| Confavreux 2014 | TOWER | Teriflunomide 14 mg PO | 370 | 43[b] | 62[b] | 43 |
| Confavreux 2014 | TOWER | Placebo | 388 | 58[b] | 86[b] | 58 |
| Fox 2012 | CONFIRM | Dimethyl fumarate 240 mg PO | 359 | 28[c] | 46[c] | 28 |
| Fox 2012 | CONFIRM | Glatiramer 20 mg SC | 350 | 38[c] | 55[c] | 38 |
| Fox 2012 | CONFIRM | Placebo | 363 | 45[c] | 61[c] | 45 |
| Gold 2012 | DEFINE | Dimethyl fumarate 240 mg PO | 409 | 52[d] | 65 | 52 |
| Gold 2012 | DEFINE | Placebo | 408 | 69[d] | 110 | 45 |
| Polman 2006 | AFFIRM | Natalizumab 300 mg IV | 627 | 69[e] | 107 | 69 |
| Polman 2006 | AFFIRM | Placebo | 315 | 72[e] | 91 | 72 |
| Coles 2008 | CAMMS223 | Alemtuzumab 12 mg IV | 112 | 8 | 16 | 8 |
| Coles 2008 | CAMMS223 | IFN B-1a 44 mcg SC | 111 | 24 | 30 | 24 |
| Cohen 2012 | CARE-MS I | Alemtuzumab 12 mg IV | 376 | 30 | | 30 |
| Cohen 2012 | CARE-MS I | IFN B-1a 44 mcg SC | 187 | 20 | | 20 |
| Coles 2012 | CARE-MS II | Alemtuzumab 12 mg IV | 426 | 54 | | 54 |
| Coles 2012 | CARE-MS II | IFN B-1a 44 mcg SC | 202 | 40 | | 40 |
| Hauser 2017 | OPERA I | Ocrelizumab 600 mg IV | 410 | 27 | 34 | 27 |
| Hauser 2017 | OPERA I | IFN B-1a 44 mcg SC | 411 | 43 | 53 | 43 |
| Hauser 2017 | OPERA II | Ocrelizumab 600 mg IV | 417 | 36 | 47 | 36 |
| Hauser 2017 | OPERA II | IFN B-1a 44 mcg SC | 418 | 56 | 73 | 56 |
| Giovannoni 2010/2011 | CLARITY | Placebo | 437 | 56[f] | 90 | 56 |
| Giovannoni 2010/2011 | CLARITY | Cladribine 3.5 mg | 433 | 35[f] | 62 | 35 |
| Giovannoni 2010/2011 | CLARITY | Cladribine 5.25 mg | 456 | 49[f] | 69 | 49 |

| Reference | Study name | Treatments | Sample size | EDSS progression 24 weeks | EDSS progression 12 weeks | EDSS progression included in NMA* |
|---|---|---|---|---|---|---|
| Kappos 2021 | OPTIMUM | Ponesimod 20 mg | 567 | 46 | 57 | 46 |
| Kappos 2021 | OPTIMUM | Teriflunomide 14 mg PO | 566 | 56 | 70 | 56 |
| Cohen 2019/Comi 2019 | RADIANCE/SUNBEAM | IFN B-1a 30 mcg IM | 889 | 36 | 69 | 36 |
| Cohen 2019/Comi 2019 | RADIANCE/SUNBEAM | Ozanimod 0.5 mg | 890 | 43 | 58 | 43 |
| Cohen 2019/Comi 2019 | RADIANCE/SUNBEAM | Ozanimod 1 mg | 880 | 51 | 67 | 51 |
| Hauser 2020 | ASCLEPIOS I | Ofatumumab | 465 | 35 | 45 | 35 |
| Hauser 2020 | ASCLEPIOS I | Teriflunomide | 459 | 53 | 63 | 53 |
| Hauser 2020 | ASCLEPIOS II | Ofatumumab | 479 | 36 | 43 | 36 |
| Hauser 2020 | ASCLEPIOS II | Teriflunomide | 472 | 46 | 62 | 46 |
| Steinman 2022 | ULTIMATE I | Teriflunomide 14 mg PO | 274 | 13 | 16 | 13 |
| Steinman 2022 | ULTIMATE I | Ublituximab IV | 271 | 9 | 14 | 9 |
| Steinman 2022 | ULTIMATE II | Teriflunomide 14 mg PO | 272 | 13 | 16 | 13 |
| Steinman 2022 | ULTIMATE II | Ublituximab IV | 272 | 9 | 14 | 9 |
| Svenningsson 2022 | RIFUND-MS | Rituximab 1000 mg IV | 98 | 10 | | 10 |
| Svenningsson 2022 | RIFUND-MS | Dimethyl fumarate 240 mg PO | 97 | 5 | | 5 |

[a] Data were obtained from the summary of product characteristics of Plegridy[25] because 24-week CDP was not reported in the pivotal publication of the trial.[26]

[b] Data were obtained from the EMA assessment report of Abaugio[27] because 24-week CDP was not reported in the pivotal publication of the trial.[28]

[c] Data were obtained from the appendix of the pivotal publication of the trial (Table S2.3 proportion of patients with EDSS progression multiplied with the sample size).[29]

[d] Data were obtained from the summary of product characteristics of Tecfidera[30] because 24-week CDP was not reported in the pivotal publication of the trial.[31]

[e] Data were obtained from the summary of product characteristics of Tysabri[32] because 24-week CDP was not reported in the pivotal publication of the trial.[33]

[f] Data were obtained from the post-hoc study of the CLARITY trial (Table 2.2: total patients – patients progression free).[34]

**Table S2.3 References RCTs included in NMA or sensitivity analysis of NMA**

| |
|---|
| Biogen Netherlands B.V. *Summary of Product Characteristics Tysabri*.; 2009. |
| Biogen Netherlands B.V. *Summary of Product Characteristics Tecfidera*.; 2013. |
| Biogen Netherlands B.V. *Summary of Product Characteristics Plegridy*.; 2014. |
| Bornstein MB, Miller A, Slagle S, et al. A pilot trial of Cop 1 in exacerbating-remitting multiple sclerosis. *The New England journal of medicine.* 1987;317(7):408-414. |
| Cadavid D, Wolansky LJ, Skurnick J, et al. Efficacy of treatment of MS with IFNbeta-1b or glatiramer acetate by monthly brain MRI in the BECOME study. *Neurology.* 2009;72(23):1976-1983. |
| Calabrese M, Bernardi V, Atzori M, et al. Effect of disease-modifying drugs on cortical lesions and atrophy in relapsing-remitting multiple sclerosis. *Mult Scler.* 2012;18(4):418-424. |
| Calabresi PA, Kieseier BC, Arnold DL, et al. Pegylated interferon beta-1a for relapsing-remitting multiple sclerosis (ADVANCE): a randomised, phase 3, double-blind study. *The Lancet Neurology.* 2014;13(7):657-665. |
| Calabresi PA, Radue EW, Goodin D, et al. Safety and efficacy of fingolimod in patients with relapsing-remitting multiple sclerosis (FREEDOMS II): a double-blind, randomised, placebo-controlled, phase 3 trial. *The Lancet Neurology.* 2014;13(6):545-556. |
| Cohen JA, Barkhof F, Comi G, et al. Oral fingolimod or intramuscular interferon for relapsing multiple sclerosis. *The New England journal of medicine.* 2010;362(5):402-415. |
| Cohen JA, Coles AJ, Arnold DL, et al. Alemtuzumab versus interferon beta 1a as first-line treatment for patients with relapsing-remitting multiple sclerosis: a randomised controlled phase 3 trial. *Lancet (London, England).* 2012;380(9856):1819-1828. |
| Cohen JA, Comi G, Selmaj KW, et al. Safety and efficacy of ozanimod versus interferon beta-1a in relapsing multiple sclerosis (RADIANCE): a multicentre, randomised, 24-month, phase 3 trial. *Lancet Neurol*. 2019;18(11):1021-1033. doi:10.1016/S1474-4422(19)30238-8 |
| Coles AJ, Compston DA, Selmaj KW, et al. Alemtuzumab vs. interferon beta-1a in early multiple sclerosis. *The New England journal of medicine.* 2008;359(17):1786-1801. |
| Coles AJ, Twyman CL, Arnold DL, et al. Alemtuzumab for patients with relapsing multiple sclerosis after disease-modifying therapy: a randomised controlled phase 3 trial. *Lancet (London, England).* 2012;380(9856):1829-1839. |
| Comi G, Kappos L, Selmaj KW, et al. Safety and efficacy of ozanimod versus interferon beta-1a in relapsing multiple sclerosis (SUNBEAM): a multicentre, randomised, minimum 12-month, phase 3 trial. *The Lancet Neurology*. 2019;18(11):1009-1020. doi:10.1016/S1474-4422(19)30239-X |
| Confavreux C, O'Connor P, Comi G, et al. Oral teriflunomide for patients with relapsing multiple sclerosis (TOWER): a randomised, double-blind, placebo-controlled, phase 3 trial. *The Lancet Neurology.* 2014;13(3):247-256. |
| Durelli L, Verdun E, Barbero P, et al. Every-other-day interferon beta-1b versus once-weekly interferon beta-1a for multiple sclerosis: results of a 2-year prospective randomised multicentre study (INCOMIN). *Lancet (London, England).* 2002;359(9316):1453-1460. |
| European Medicines Agency. *Assessment Report Aubagio*.; 2013. |
| Etemadifar M, Janghorbani M, Shaygannejad V. Comparison of Betaferon, Avonex, and Rebif in treatment of relapsing-remitting multiple sclerosis. *Acta neurologica Scandinavica.* 2006;113(5):283-287. |
| Fox RJ, Miller DH, Phillips JT, et al. Placebo-controlled phase 3 study of oral BG-12 or glatiramer in multiple sclerosis. *The New England journal of medicine.* 2012;367(12):1087-1097. |
| Giovannoni G, Comi G, Cook S, et al. A Placebo-Controlled Trial of Oral Cladribine for Relapsing Multiple Sclerosis. New England Journal of Medicine. 2010;362(5):416-426. doi:10.1056/NEJMoa0902533 |
| Giovannoni G, Cook S, Rammohan K, et al. Sustained disease-activity-free status in patients with relapsing-remitting multiple sclerosis treated with cladribine tablets in the CLARITY study: a post-hoc and subgroup analysis. *Lancet Neurol*. 2011;10(4):329-337. doi:10.1016/S1474-4422(11)70023-0 |

Gold R, Giovannoni G, Selmaj K, et al. Daclizumab high-yield process in relapsing-remitting multiple sclerosis (SELECT): a randomised, double-blind, placebo-controlled trial. *Lancet (London, England).* 2013;381(9884):2167-2175.

Gold R, Kappos L, Arnold DL, et al. Placebo-controlled phase 3 study of oral BG-12 for relapsing multiple sclerosis. *The New England journal of medicine.* 2012;367(12):1098-1107.

Hauser SL, Bar-Or A, Comi G, et al. Ocrelizumab versus Interferon Beta-1a in Relapsing Multiple Sclerosis. *New England Journal of Medicine.* 2017;376(3):221-234.

Hauser SL, Waubant E, Arnold DL, et al. B-cell depletion with rituximab in relapsing-remitting multiple sclerosis. *The New England journal of medicine.* 2008;358(7):676-688.

Hauser SL, Bar-Or A, Cohen JA, et al. Ofatumumab versus Teriflunomide in Multiple Sclerosis. *New England Journal of Medicine*. 2020;383(6):546-557. doi:10.1056/NEJMoa1917246

Interferon beta-1b is effective in relapsing-remitting multiple sclerosis. I. Clinical results of a multicenter, randomized, double-blind, placebo-controlled trial. The IFNB Multiple Sclerosis Study Group. *Neurology.* 1993;43(4):655-661.

Jacobs LD, Cookfair DL, Rudick RA, et al. Intramuscular interferon beta-1a for disease progression in relapsing multiple sclerosis. The Multiple Sclerosis Collaborative Research Group (MSCRG). *Annals of neurology.* 1996;39(3):285-294.

Johnson KP, Brooks BR, Cohen JA, et al. Copolymer 1 reduces relapse rate and improves disability in relapsing-remitting multiple sclerosis: results of a phase III multicenter, double-blind placebo-controlled trial. The Copolymer 1 Multiple Sclerosis Study Group. *Neurology.* 1995;45(7):1268-1276.

Kappos L, Radue EW, O'Connor P, et al. A placebo-controlled trial of oral fingolimod in relapsing multiple sclerosis. *The New England journal of medicine.* 2010;362(5):387-401.

Kappos L, Wiendl H, Selmaj K, et al. Daclizumab HYP versus Interferon Beta-1a in Relapsing Multiple Sclerosis. *The New England journal of medicine.* 2015;373(15):1418-1428.

Kappos L, Fox RJ, Burcklen M, et al. Ponesimod Compared With Teriflunomide in Patients With Relapsing Multiple Sclerosis in the Active-Comparator Phase 3 OPTIMUM Study: A Randomized Clinical Trial. *JAMA Neurol*. 2021;78(5):558-567. doi:10.1001/jamaneurol.2021.0405

Khan O, Rieckmann P, Boyko A, Selmaj K, Zivadinov R. Three times weekly glatiramer acetate in relapsing-remitting multiple sclerosis. Annals of neurology. 2013;73(6):705-713.

Lublin FD, Cofield SS, Cutter GR, et al. Randomized study combining interferon and glatiramer acetate in multiple sclerosis. *Annals of neurology.* 2013;73(3):327-340.

Mikol DD, Barkhof F, Chang P, et al. Comparison of subcutaneous interferon beta-1a with glatiramer acetate in patients with relapsing multiple sclerosis (the REbif vs Glatiramer Acetate in Relapsing MS Disease [REGARD] study): a multicentre, randomised, parallel, open-label trial. *The Lancet Neurology.* 2008;7(10):903-914.

O'Connor P, Filippi M, Arnason B, et al. 250 microg or 500 microg interferon beta-1b versus 20 mg glatiramer acetate in relapsing-remitting multiple sclerosis: a prospective, randomised, multicentre study. *The Lancet Neurology.* 2009;8(10):889-897.

O'Connor P, Wolinsky JS, Confavreux C, et al. Randomized trial of oral teriflunomide for relapsing multiple sclerosis. *The New England journal of medicine.* 2011;365(14):1293-1303.

Panitch H, Goodin DS, Francis G, et al. Randomized, comparative study of interferon beta-1a treatment regimens in MS: The EVIDENCE Trial. *Neurology.* 2002;59(10):1496-1506.

Polman CH, O'Connor PW, Havrdova E, et al. A randomized, placebo-controlled trial of natalizumab for relapsing multiple sclerosis. *The New England journal of medicine.* 2006;354(9):899-910.

Randomised double-blind placebo-controlled study of interferon beta-1a in relapsing/remitting multiple sclerosis. PRISMS (Prevention of Relapses and Disability by Interferon beta-1a Subcutaneously in Multiple Sclerosis) Study Group. *Lancet (London, England).* 1998;352(9139):1498-1504.

| |
|---|
| Svenningsson A, Frisell T, Burman J, et al. Safety and efficacy of rituximab versus dimethyl fumarate in patients with relapsing-remitting multiple sclerosis or clinically isolated syndrome in Sweden: a rater-blinded, phase 3, randomised controlled trial. *The Lancet Neurology*. 2022;21(8):693-703. doi:10.1016/S1474-4422(22)00209-5 |
| Vermersch P, Czlonkowska A, Grimaldi LM, et al. Teriflunomide versus subcutaneous interferon beta-1a in patients with relapsing multiple sclerosis: a randomised, controlled phase 3 trial. *Mult Scler.* 2014;20(6):705-716. |
| Vollmer TL, Sorensen PS, Selmaj K, et al. A randomized placebo-controlled phase III trial of oral laquinimod for multiple sclerosis. *J Neurol.* 2014;261(4):773-783. |

### Results

#### Included studies

References of the included studies can be found in Table S2.3.

#### Relapses

The NMA for annualized relapse rates (ARR) was conducted with data from 35 RCTs, with a total sample size of 26,096 patients in which a total of 12,977 events were observed. The outcome statistic is the incidence rate ratio as captured by the number of events by person years using the DMT relative to placebo. The network is centred around placebo-controlled trials, as can be seen from Figure S2.1. Figure S2.2 provides the incidence rate ratios (IRR) and their confidence intervals for the included DMTs derived from the network met-analysis.

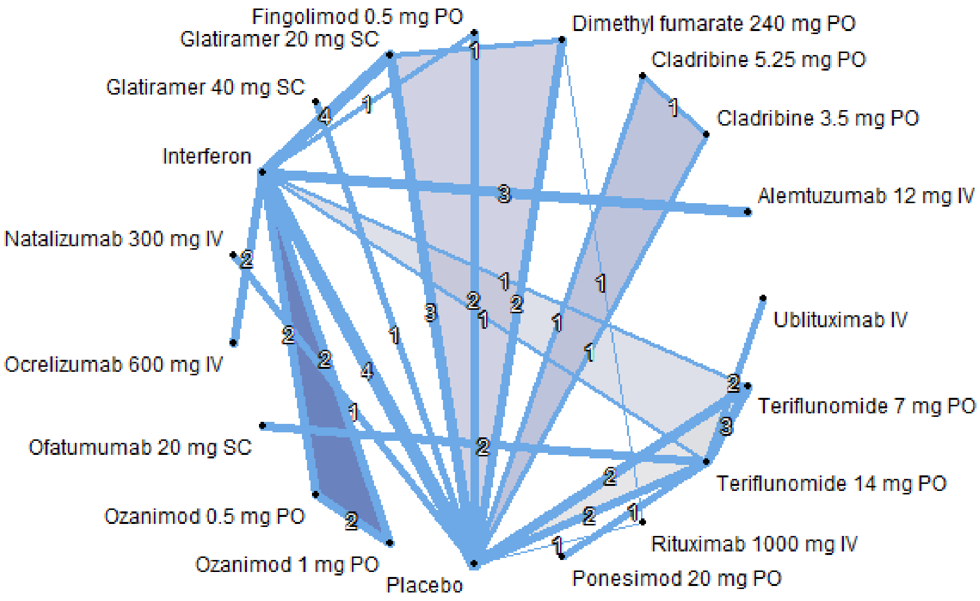

**Figure S2.1 Network graph of annualized relapse rates.** Shaded areas represent RCTs with three arms.

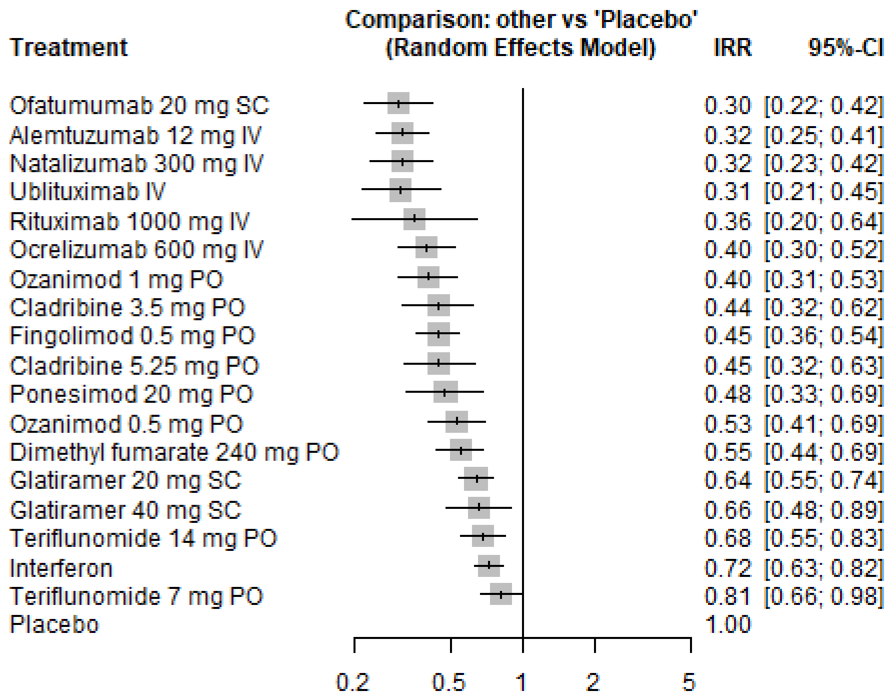

**Figure S2.2** Results network-meta analysis incidence rate ratio (IRR) of the annualized relapse rate (ARR, $I^2$=53.5%)

Disability progression

The NMA for disability progression as captured by EDSS score was conducted with data from 25 trials, with a total sample size of 20,433 patients in which for 2,219 patients 24-week CDP was observed. The outcome statistic is a relative risk (RR) on disability progression with a DMT relative to placebo. The network is centred around placebo-controlled trials, as can be seen from Figure S2.3. Figure S2.4 provides the relative risks and their confidence intervals for the included DMTs derived from the network meta-analysis.

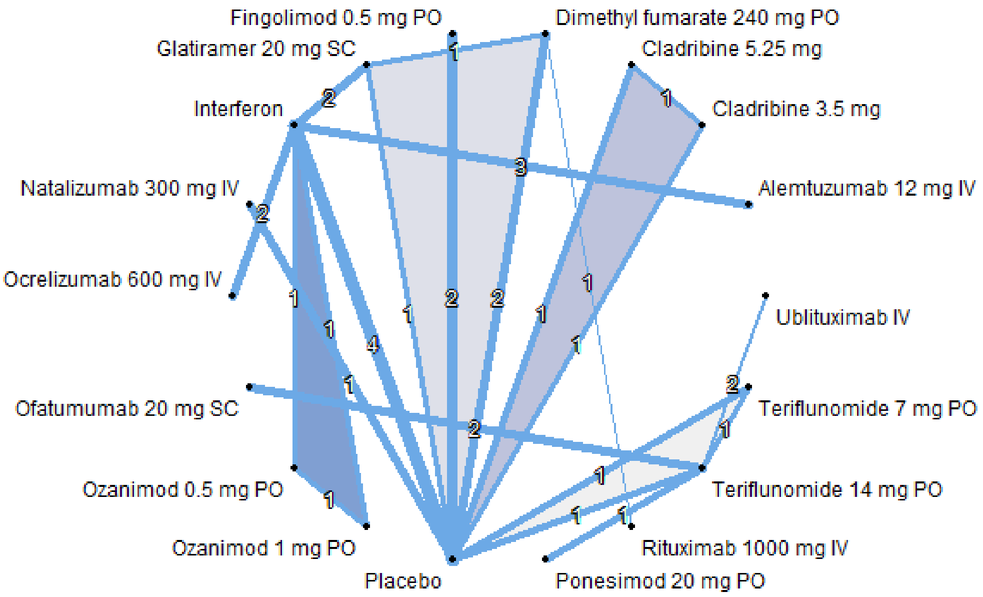

**Figure S2.3 Network graph of disability progression.** Shaded areas represent RCTs with three arms.

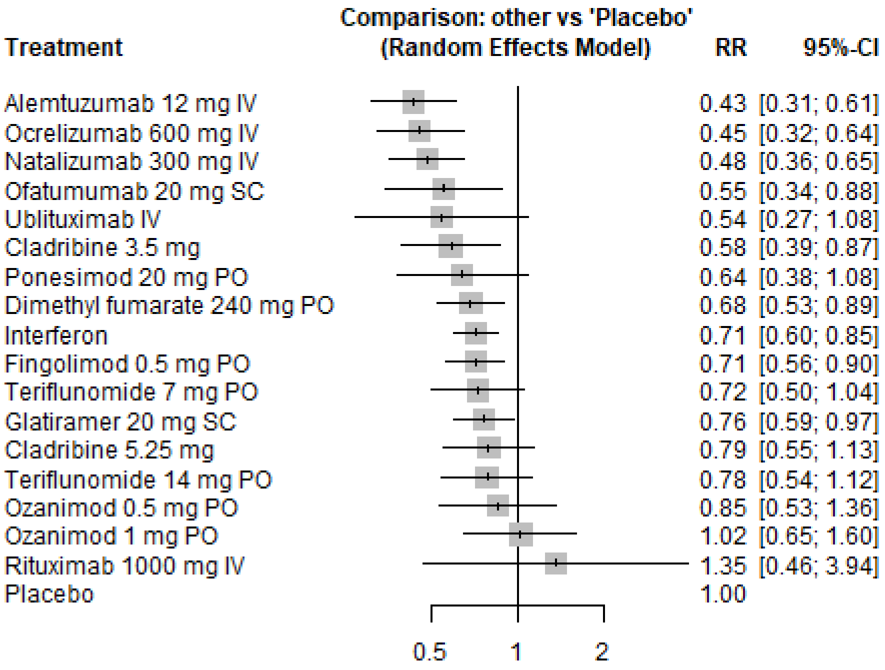

**Figure S2.4** Results network-meta analysis relative risk of disability progression (EDSS, $I^2$=0%)

*Sensitivity analysis with clustered S1PR-modulators*

We performed a sensitivity analysis clustering the S1PR-modulators into one class: fingolimod, ponesimod, and ozanimod. The results are shown in Figure S2.5. The differences between the base-case analysis with individual S1PR-modulators are minor.

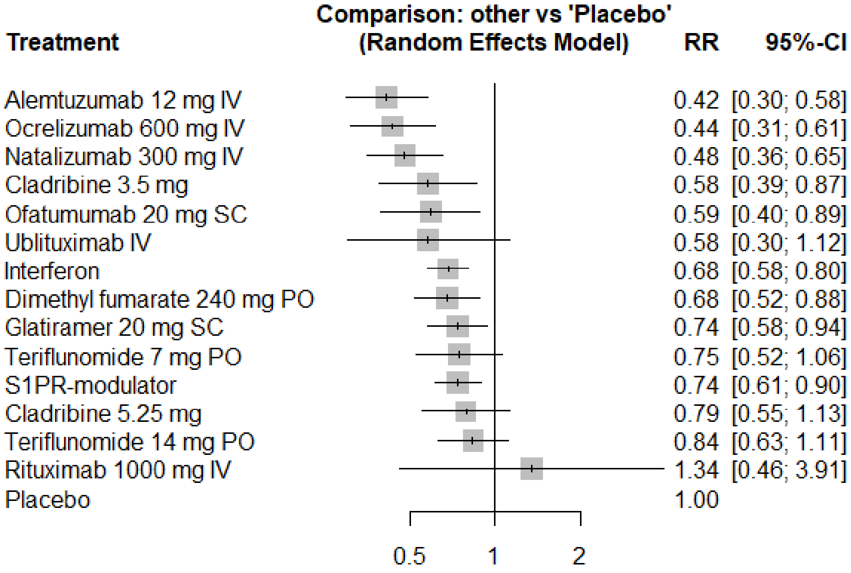

**Figure S2.5 Results network-meta analysis relative risk of disability progression with clustered S1PR-modulators (EDSS, $I^2$=0%)**

*Sensitivity analysis 24-week CDP versus 12- and 24-week CDP*

We performed a sensitivity analysis including the five studies that only reported 12-week confirmed disease progression (CDP) instead of 24-week CDP. The results are shown in Table S2.4 and Figure S2.5. The differences between the base-case analysis that only included studies with 24-week CDP are minor, except for the inclusion of glatiramer 40 mg because there was only a study that reported 12-week CDP for this DMT.

**Table S2.4 Results network-meta analysis relative risk of disability progression (EDSS) including studies that reported only 12-week CDP instead of 24-week CDP.**

| | Only 24 week CDP (base-case) | | Both 12- and 24 week CDP | | Difference | |
|---|---|---|---|---|---|---|
| Treatment | RR | seRR | RR | seRR | ΔRR | ΔseRR |
| Alemtuzumab 12 mg IV | 0.433 | 0.171 | 0.457 | 0.166 | -0.023 | -0.005 |
| Cladribine 3.5 mg | 0.582 | 0.203 | 0.582 | 0.203 | 0.000 | 0.000 |
| Cladribine 5.25 mg | 0.789 | 0.182 | 0.789 | 0.182 | 0.000 | 0.000 |
| Dimethyl fumarate 240 mg PO | 0.683 | 0.134 | 0.684 | 0.133 | -0.001 | -0.001 |
| Fingolimod 0.5 mg PO | 0.710 | 0.120 | 0.679 | 0.109 | 0.031 | -0.010 |
| Glatiramer 20 mg SC | 0.760 | 0.126 | 0.767 | 0.096 | -0.006 | -0.029 |
| Glatiramer 40 mg SC | | | 1.208 | 0.282 | | |
| Interferons | 0.712 | 0.090 | 0.750 | 0.081 | -0.038 | -0.009 |
| Natalizumab 300 mg IV | 0.481 | 0.154 | 0.481 | 0.154 | 0.000 | 0.000 |
| Ocrelizumab 600 mg IV | 0.454 | 0.178 | 0.479 | 0.174 | -0.024 | -0.004 |
| Ofatumumab 20 mg SC | 0.550 | 0.239 | 0.530 | 0.189 | 0.020 | -0.050 |
| Ozanimod 0.5 mg PO | 0.850 | 0.239 | 0.895 | 0.235 | -0.045 | -0.003 |
| Ozanimod 1 mg PO | 1.019 | 0.231 | 1.074 | 0.227 | -0.054 | -0.003 |
| Ponesimod 20 mg PO | 0.637 | 0.267 | 0.614 | 0.223 | 0.023 | -0.044 |
| Rituximab 1000 mg IV | 1.351 | 0.545 | 1.353 | 0.545 | | |
| Teriflunomide 14 mg PO | 0.777 | 0.188 | 0.749 | 0.117 | 0.028 | -0.071 |
| Teriflunomide 7 mg PO | 0.723 | 0.187 | 0.762 | 0.114 | -0.039 | -0.073 |
| Ublituximab IV | 0.541 | 0.354 | 0.521 | 0.322 | 0.020 | -0.032 |

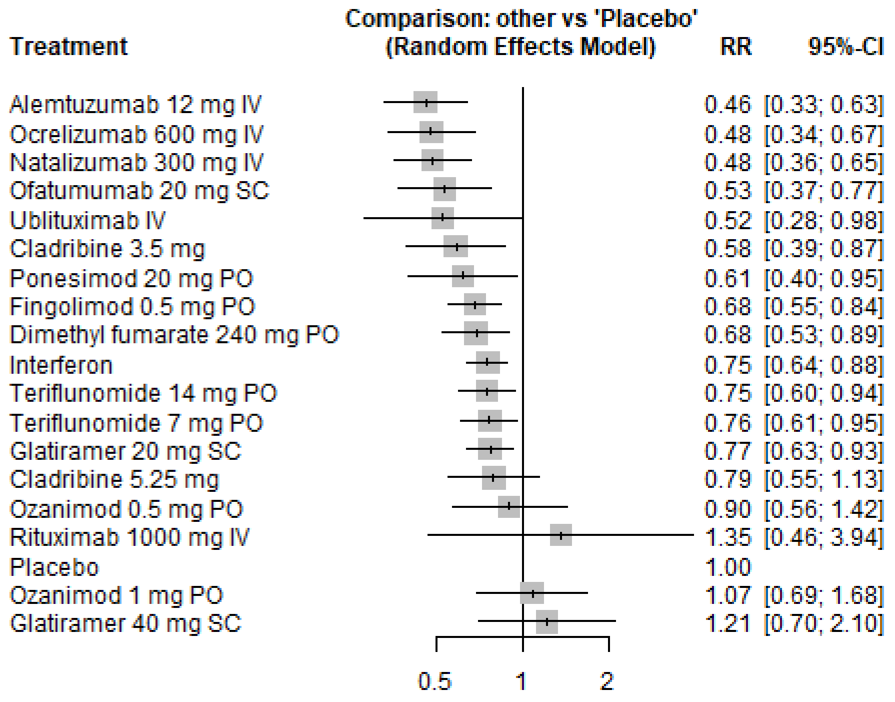

**Figure S2.6. Results network-meta analysis relative risk of disability progression (EDSS) including studies that reported only 12-week CDP instead of 24-week CDP.**

Between-study heterogenenity

The I-squared statistic was 53.5% and 0% for relapse and disability progression, respectively. These results suggest limited moderate and no between-study heterogeneity for relapse and disability progression, respectively.

Direct and indirect evidence

Direct and indirect evidence compared well, with no significant differences as shown in Figure S2.6 and S2.7 for relapse and disability progression respectively.

| Comparison | Number of Studies | Direct Evidence | I2 | IRR | 95%-CI |
|---|---|---|---|---|---|
| **Dimethyl fumarate 240 mg PO:Glatiramer 20 mg SC** | | | | | |
| Direct estimate | 1 | 0.52 | | 0.78 | [0.56; 1.10] |
| Indirect estimate | | | | 0.96 | [0.67; 1.37] |
| Network estimate | | | | 0.86 | [0.67; 1.10] |
| **Dimethyl fumarate 240 mg PO:Placebo** | | | | | |
| Direct estimate | 2 | 0.89 | 0.02 | 0.55 | [0.43; 0.69] |
| Indirect estimate | | | | 0.56 | [0.28; 1.10] |
| Network estimate | | | | 0.55 | [0.44; 0.69] |
| **Dimethyl fumarate 240 mg PO:Rituximab 1000 mg IV** | | | | | |
| Direct estimate | 1 | 0.24 | | 5.39 | [1.53; 18.97] |
| Indirect estimate | | | | 1.04 | [0.51; 2.11] |
| Network estimate | | | | 1.54 | [0.83; 2.85] |
| **Fingolimod 0.5 mg PO:Interferon** | | | | | |
| Direct estimate | 1 | 0.32 | | 0.49 | [0.33; 0.71] |
| Indirect estimate | | | | 0.69 | [0.53; 0.90] |
| Network estimate | | | | 0.62 | [0.50; 0.77] |
| **Fingolimod 0.5 mg PO:Placebo** | | | | | |
| Direct estimate | 2 | 0.76 | 0.09 | 0.48 | [0.39; 0.61] |
| Indirect estimate | | | | 0.34 | [0.23; 0.51] |
| Network estimate | | | | 0.45 | [0.36; 0.54] |
| **Glatiramer 20 mg SC:Interferon** | | | | | |
| Direct estimate | 4 | 0.67 | 0.33 | 0.88 | [0.74; 1.05] |
| Indirect estimate | | | | 0.88 | [0.69; 1.14] |
| Network estimate | | | | 0.88 | [0.76; 1.02] |
| **Glatiramer 20 mg SC:Placebo** | | | | | |
| Direct estimate | 3 | 0.52 | 0.86 | 0.64 | [0.51; 0.79] |
| Indirect estimate | | | | 0.64 | [0.51; 0.80] |
| Network estimate | | | | 0.64 | [0.55; 0.74] |
| **Interferon:Placebo** | | | | | |
| Direct estimate | 4 | 0.64 | 0.20 | 0.72 | [0.61; 0.85] |
| Indirect estimate | | | | 0.72 | [0.58; 0.89] |
| Network estimate | | | | 0.72 | [0.63; 0.82] |
| **Interferon:Teriflunomide 14 mg PO** | | | | | |
| Direct estimate | 1 | 0.17 | | 0.84 | [0.48; 1.49] |
| Indirect estimate | | | | 1.11 | [0.86; 1.43] |
| Network estimate | | | | 1.06 | [0.84; 1.34] |
| **Interferon:Teriflunomide 7 mg PO** | | | | | |
| Direct estimate | 1 | 0.18 | | 0.52 | [0.31; 0.89] |
| Indirect estimate | | | | 1.01 | [0.78; 1.29] |
| Network estimate | | | | 0.89 | [0.71; 1.12] |
| **Rituximab 1000 mg IV:Placebo** | | | | | |
| Direct estimate | 1 | 0.78 | | 0.51 | [0.26; 0.99] |
| Indirect estimate | | | | 0.10 | [0.03; 0.35] |
| Network estimate | | | | 0.36 | [0.20; 0.64] |
| **Teriflunomide 14 mg PO:Placebo** | | | | | |
| Direct estimate | 2 | 0.88 | 0.00 | 0.66 | [0.53; 0.82] |
| Indirect estimate | | | | 0.80 | [0.45; 1.43] |
| Network estimate | | | | 0.68 | [0.55; 0.83] |
| **Teriflunomide 7 mg PO:Placebo** | | | | | |
| Direct estimate | 2 | 0.86 | 0.15 | 0.74 | [0.59; 0.91] |
| Indirect estimate | | | | 1.43 | [0.84; 2.44] |
| Network estimate | | | | 0.81 | [0.66; 0.98] |
| **Teriflunomide 14 mg PO:Teriflunomide 7 mg PO** | | | | | |
| Direct estimate | 3 | 1.00 | 0.63 | 0.85 | [0.69; 1.04] |
| Indirect estimate | | | | 0.00 | [0.00; 2.07] |
| Network estimate | | | | 0.84 | [0.69; 1.03] |

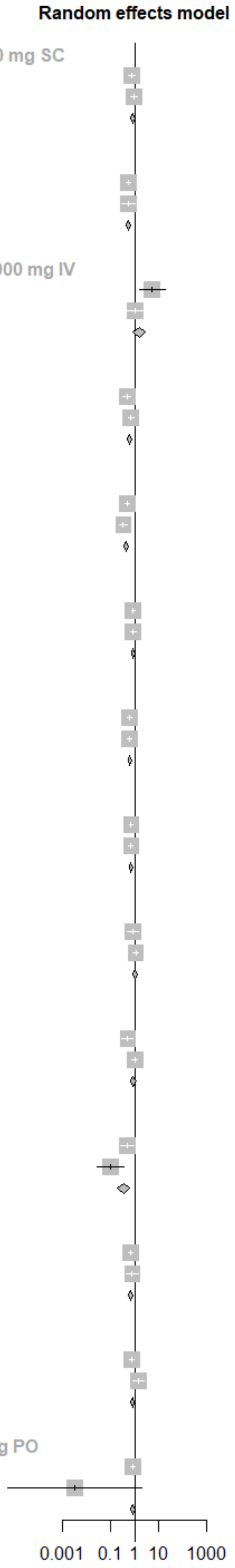

**Figure S2.6 Comparison of direct and indirect evidence for ARR**

| Comparison | Number of Studies | Direct Evidence | I2 | RR | 95%-CI |
|---|---|---|---|---|---|
| **Dimethyl fumarate 240 mg PO:Glatiramer 20 mg SC** | | | | | |
| Direct estimate | 1 | 0.50 | | 0.72 | [0.45; 1.14] |
| Indirect estimate | | | | 1.12 | [0.70; 1.77] |
| Network estimate | | | | 0.90 | [0.65; 1.25] |
| **Dimethyl fumarate 240 mg PO:Placebo** | | | | | |
| Direct estimate | 2 | 0.96 | 0.00 | 0.71 | [0.54; 0.92] |
| Indirect estimate | | | | 0.30 | [0.08; 1.14] |
| Network estimate | | | | 0.68 | [0.53; 0.89] |
| **Glatiramer 20 mg SC:Interferon** | | | | | |
| Direct estimate | 2 | 0.73 | 0.59 | 0.98 | [0.76; 1.27] |
| Indirect estimate | | | | 1.34 | [0.88; 2.06] |
| Network estimate | | | | 1.07 | [0.86; 1.33] |
| **Glatiramer 20 mg SC:Placebo** | | | | | |
| Direct estimate | 1 | 0.37 | | 0.88 | [0.58; 1.31] |
| Indirect estimate | | | | 0.70 | [0.51; 0.95] |
| Network estimate | | | | 0.76 | [0.59; 0.97] |
| **Interferon:Placebo** | | | | | |
| Direct estimate | 4 | 0.85 | 0.04 | 0.68 | [0.56; 0.82] |
| Indirect estimate | | | | 0.93 | [0.59; 1.48] |
| Network estimate | | | | 0.71 | [0.60; 0.85] |

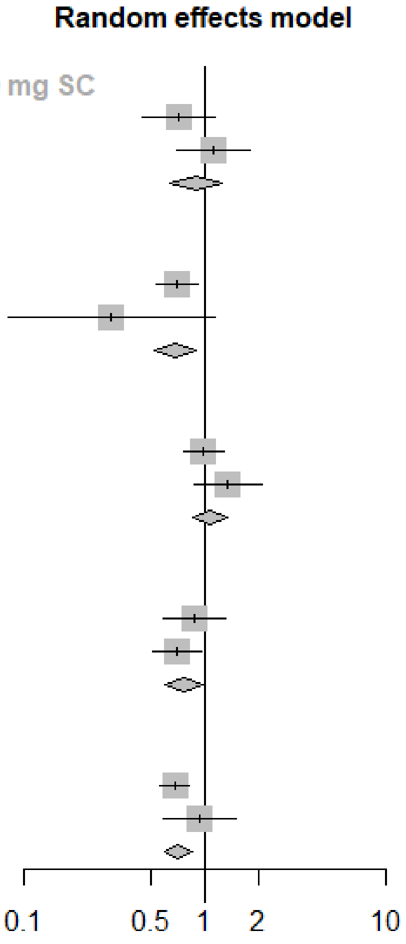

**Figure S7: Comparison of direct and indirect evidence for EDSS progression**
